# The effectiveness of acupoint herbal patching for non-specific low back pain: A Protocol for systematic review and meta-analysis

**DOI:** 10.1101/2022.01.25.22269813

**Authors:** Tianshu Ma, WU Liu, Yan Dong, Lei Cheng, Ziyuan Wang, Xiaona Liu, Tie Li, Chengyu Liu

## Abstract

**Background:** Non-specific low back pain(NS-LBP) is related to disability and work absence and accounts for high economical costs of global.It is a problem that has a negative impact on physical, mental health of patients and quality of life. At present, acupoint herbal patching(AHP) as an adjuvant therapy is currently undergoing clinical trials in different medical centers. This study aimed to design a systematic review and meta-analysis to explore the effects of AHP on non-specific low back pain(NS-LBP).

**Methods:** We will search the Cochrane Central Register of Controlled Trials, PubMed, Embase, the Web of Science, the Chinese Biomedical Literature Database,the Chinese Scientific Journal Database, the Wan-Fang Database and the China National Knowledge Infrastructure. The primary outcome measures will be clinical effective rate, functional outcomes, and quality of life. Data that meets the inclusion criteria will be extracted and analyzed using RevMan V.5.3 software. Two reviewers will evaluate the studies using the Cochrane Collaboration risk of bias tool. We will use the GRADE approach to assess the overall quality of evidence supporting the primary outcomes. We will also use Spass software (Version19.0) for complex network analysis to explore the potential core prescription of acupoint herbal patching for functional constipation.

**Results:** This systematic review protocol will analyze the effectiveness, quality of life, improvement of the symptom and safety of acupoint herbal patching therapy for non-specific low back pain.

**Conclusion:** The findings of this systematic review will provide evidence to evaluate the effectiveness and safety of acupoint herbal patching for non-specific low back pain.

## 1. Introduction

Non-specific low back pain(NS-LBP)is defined as low back pain not attributable to a recognisable, known specific pathology(eg, infection, tumour, osteoporosis, fracture, structural deformity, infl ammatory disorder, radicular syndrome, or cauda equina syndrome). Low back pains to became one of the biggest problems of public health systems in the western world during the second half of the 20th century, and now seems to be extending worldwide[1,2].Data onto the USA show that the proportion of physician visits attributed to back pain has changed little into the past decade[3],but the cost has increased substantially[4].Findings from a UK survey showed that the annual consultation prevalence for low back pain was 417 per 10000 registered patients. The lowest rate was recorded in the 0-14year age-group (30 per 10000) and the highest in the 45-64 year age-group (536 per 10000)[5].

Non-specific LBP has become a major public health problem worldwide. The lifetime prevalence of low back pain is reported to be as high as 84%, and the prevalence of chronic low back pain is about 23%, with 11-12% of the population being disabled by low back pain[6]. The management of LBP comprises to arange of different intervention strategies including surgery, non-medical interventions such as exercise, behavioral therapy, and alternative therapies. Pharmacological interventions are the most frequently recommended intervention in back pain [7-8].Low back pain is a common reason to visit a general practitioner[9-10], when patients are often prescribed pharmacological interventions to manage their symptoms[11-13].The commonly used drug intervention includes:pharmacological interventions(i.e., non-steroid anti-inflammatory drugs (NSAIDs), muscle relaxants,antidepressants,and opioids)for LBP[14].According to the authors of the studies on NSAIDs,most adverse effects, including abdominal pain, diarrhea,edema, dry mouth, rash, dizziness, headache, and tiredness,were considered to be mild to moderately severe[15].NSAIDs and opioids might be useful for short-term pain relief in patients with chronic LBP, who responded with an exacerbation of their symptoms after stopping their medication.For antidepressants adverse effects, such as dry mouth,constipation, tachycardia,sedation,orthostatic hypotension,and tremor,were commonly reported[16].Recommendations for the use of muscle relaxants have, however, conflicted between international clinical practice guidelines for low back pain[17-19].For example,the US guideline recommends non-benzodiazepine antispasmodics as the drug of choice for acute low back pain[20],the Belgian guideline discourages such use[21],and the UK guideline does not make a recommendation[22].Therefore, for treatment of LBP can’t uniform standard.These methods have brought significant economic burdens and thus considerable healthcare utilization to patients.At the same time, LBP seriously impairs the physical and mental health of patients and affects their quality of life.Therefore, many people, including those who do not improve with existing medications or suffer many side effects, are interested in complementary and alternative medicine.

Acupoint herbal patching (AHP) is a combination treatment method of externally applying a processed herbal preparation patch to acupoints. It has the advantage that various herbs can be attached to various acupoints without serious systemic adverse events. The earliest records of AHP is found in the Prescriptions for Fifty-two Diseases (Wu Shi Er Bing Fang), and it has been widely used for preventing various diseases by strengthening the immune system through the stimulation of acupoints using various herbal preparations[23-25]. AHP is an inexpensive, easy-to-use, noninvasive, and safe treatment method, which can increase compliance with LBP patients. Modern studies suggest that AHP can penetrate Chinese medicine effective component percutaneous absorption, avoid the liver and kidney, gastrointestinal absorption of drugs directly reduces the liver and kidney damage, also won’t destroy the gastrointestinal function[26].At the same time it can adjust local blood circulation, improve immunity, and finally obtain the exact therapeutic effect.As a complementary and alternative therapy, AHP is often used to treat aches and pains diseases, and its application in LBP has gradually become popular in recent years.However,the clinical efficacy and potential treatment prescriptions of AHP for LBP remain unclear, requiring further exploration. Therefore, asystematic evaluation of treatment outcomes and treatment prescriptions may help better explain and push this method into practice. In this study, we will investigate current evidence associated with the effectiveness and safety of AHP for LBP, which will help clinicians to better use it in clinical practice.

## 2. Methods

We will collect randomized controlled trials (RCTs) to evaluate clinical effectiveness, functional outcomes, quality of life, and side effects of AHP on LBP for systematic review and meta-analysis.RCTs comparing AHP for FC with no treatment, placebo, or conventional drugs will be included.All eligible trials will be included regardless of language and publication type. RCTs that meet the requirements will be included for data mining. Articles of the following research types will be excluded: case series, observational studies (including cohort studies and case-control studies) and retrospective studies,qualitative studies, animal experiments, review articles. In addition, there will be no restrictions on study area, race, patient gender.

## 3. Inclusion and exclusion criteria

### Types of participants

In order to be included in this review, participants of the RCTs must fulfill the following inclusion criteria: adult subjects (C18 years of age) with nonspecific LBP (including discopathy or any other non-specific degenerative pathology such as osteoarthritis).

### The exclusion criteria were

1. trials including subjectswith specific LBP caused by pathologies such as vertebral spinal stenosis, ankylosing spondylitis, scoliosis, and coccydynia;
2. trials including subjectswith specific LBP caused by infection, tumour, osteoporosis, fracture, structural deformity, infl ammatory disorder, radicular syndrome, or cauda equina syndrome;
3. post-partumLBP or pelvic pain due to pregnancy;
4. post-operativestudies;
5. prevention studies;
6. abstracts or nonpublished studies.

### Types of interventions

Participants in the intervention group are those undergoing acupoint herbal patching, regardless of herbal regimen, acupoints selected, patching time. There will not be any restrictions on age and original countries of the participants. In the control group,patients received medication, no treatment, sham or placebo acupoint catgut embedding, acupuncture/electro-acupuncture and etc. The other interventions between the control group and the intervention group should be the same.

### Types of outcome measures

For inclusion, at least one of the following outcome measures should have been measured in the RCT: pain intensity [e.g., visual analog scale (VAS), numerical rating scale (NRS), McGill pain questionnaire], back-specific functional status (e.g., Roland-Morris Disability Questionnaire, Oswestry Disability Index), perceived recovery(e.g., overall improvement), and return to work (e.g., return to work status, sick leave days). The primary outcomes for this review were pain and functional status.

## 4. Search strategy

An electronic search will be conducted. We will identify relevant studies from the Cochrane Central Register of Controlled Trials,PubMed, Embase, the Web of Science, the Chinese Biomedical Literature Database (CBM), the Chinese Scientific Journal Database (CSJD), the Wan-Fang Database (Wanfang) and the China National Knowledge Infrastructure (CNKI) from their inception to 15 January 2021. The search term will consist of 3 parts: intervention method, disease, and study type: (“acupoint application” or “ acupoint sticker” or “crude herb moxibustion”or “medicinal vesiculation” or “herbal patch” or “herbal plaster” or “acupoint patch” or “Sanfu” or “acupoint sticking”or “point application therapy” or “drug acupoint application” or“winter diseases treated with acupoint stimulation in summer” or“drugs and points for point application in summer to treat the diseases with attacks in winter”or “acupuncture point application therapies” or “plaster therapy” or “external application therapy” or “acupoint herbal patching”) and(“low back pain” or“ non-specific low-back pain”or“Chronic low back pain”or“Acute low back pain”or“Subacute and chronic low back pain”or “persistent low back pain”) and (“randomized controlled trial” or “randomized” or “case control studies” or “observational studies”or “case series” or “trial”) and (“blind”).The details of the PubMed Database search strategies are provided in Tables 1. The similar but adaptive search strategies will be applied to other electronic databases. Language will be restricted to English and Chinese. Reference lists of relevant original studies will be screened to identify additional potentially citations. In addition,the following 3 trial registries will be searched for ongoing studies: Current Controlled Trials: www.controlled-trials.com;Clinical Trials: www.ClinicalTrials.gov; and Chinese Clinical Trial Registry: www.chictr.org.cn/index.aspx.

**Table 1.**
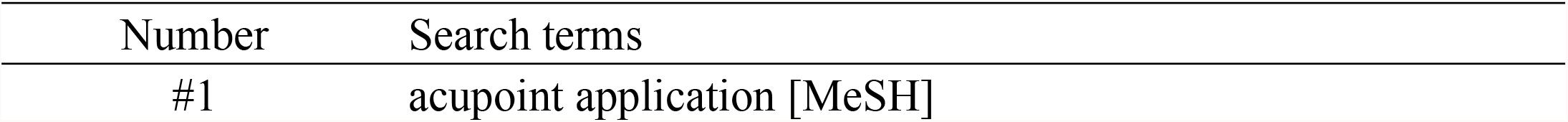
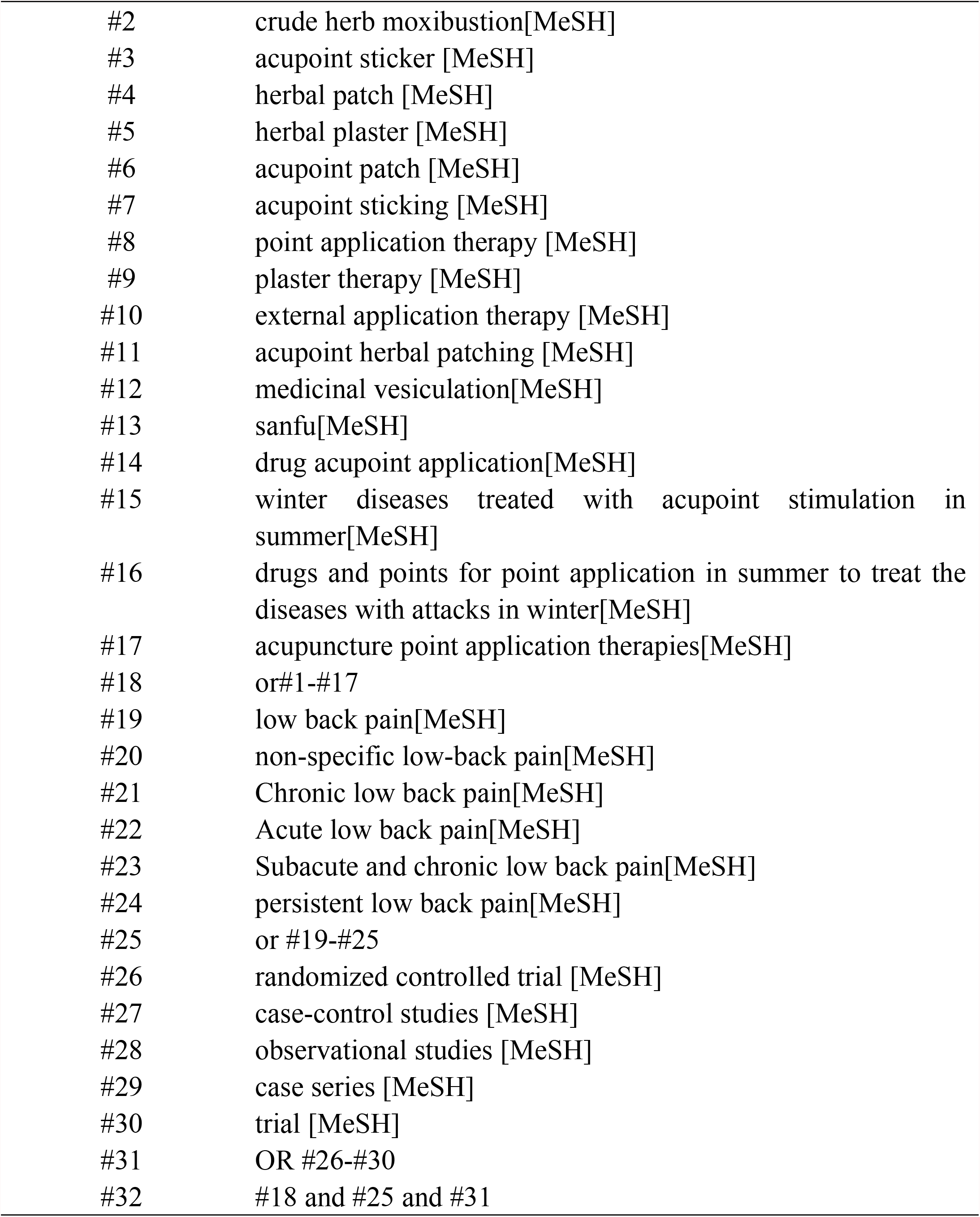
The search strategy for PubMed database

| Number | Search terms |
| --- | --- |
| #1 | acupoint application [MeSH] |
| #2 | crude herb moxibustion[MeSH] |
| #3 | acupoint sticker [MeSH] |
| #4 | herbal patch [MeSH] |
| #5 | herbal plaster [MeSH] |
| #6 | acupoint patch [MeSH] |
| #7 | acupoint sticking [MeSH] |
| #8 | point application therapy [MeSH] |
| #9 | plaster therapy [MeSH] |
| #10 | external application therapy [MeSH] |
| #11 | acupoint herbal patching [MeSH] |
| #12 | medicinal vesiculation[MeSH] |
| #13 | sanfu[MeSH] |
| #14 | drug acupoint application[MeSH] |
| #15 | winter diseases treated with acupoint stimulation in summer[MeSH] |
| #16 | drugs and points for point application in summer to treat the diseases with attacks in winter[MeSH] |
| #17 | acupuncture point application therapies[MeSH] |
| #18 | or #1-#17 |
| #19 | low back pain[MeSH] |
| #20 | non-specific low-back pain[MeSH] |
| #21 | Chronic low back pain[MeSH] |
| #22 | Acute low back pain[MeSH] |
| #23 | Subacute and chronic low back pain[MeSH] |
| #24 | persistent low back pain[MeSH] |
| #25 | or #19-#25 |
| #26 | randomized controlled trial [MeSH] |
| #27 | case-control studies [MeSH] |
| #28 | observational studies [MeSH] |
| #29 | case series [MeSH] |
| #30 | trial [MeSH] |
| #31 | OR #26-#30 |
| #32 | #18 and #25 and #31 |

## 5. Study selection and data extraction

Author (Ma TS) with experience in the field will guide the search.First, the NoteExpress 3.2.0 software (Available at: http://www.inoteexpress.com/aegean/) will be used to exclude duplicate references from different databases.Two review authors (Ma TS,Dong Y) will independently assess the title and abstracts of all citations found from the above search strategy. A copy of the full text article is obtained for the potentially eligible studies. These review authors will independently read the full text articles to include eligible studies; disagreement will be resolved by consensus through discussion with a third review author (Dong Y). If conclusion still cannot be met, we will contact the author of the article to determine the eligibility of the study. The selection process will be showed in a PRISMA flow chart (http://www.prismastatementorg/) (Fig. 1). In the end, Two review authors (Liu CY,Ma TS) will extract data using a data extraction form according to the recommendations of the Cochrane Handbook for Systematic Reviews of Interventions. the following data will be extracted: author, year of publication, country where the study was conducted, study period, original inclusion criteria, total number of people included in the study, acupoints, doses of herbs and time of application and etc.

**Figure 1.**
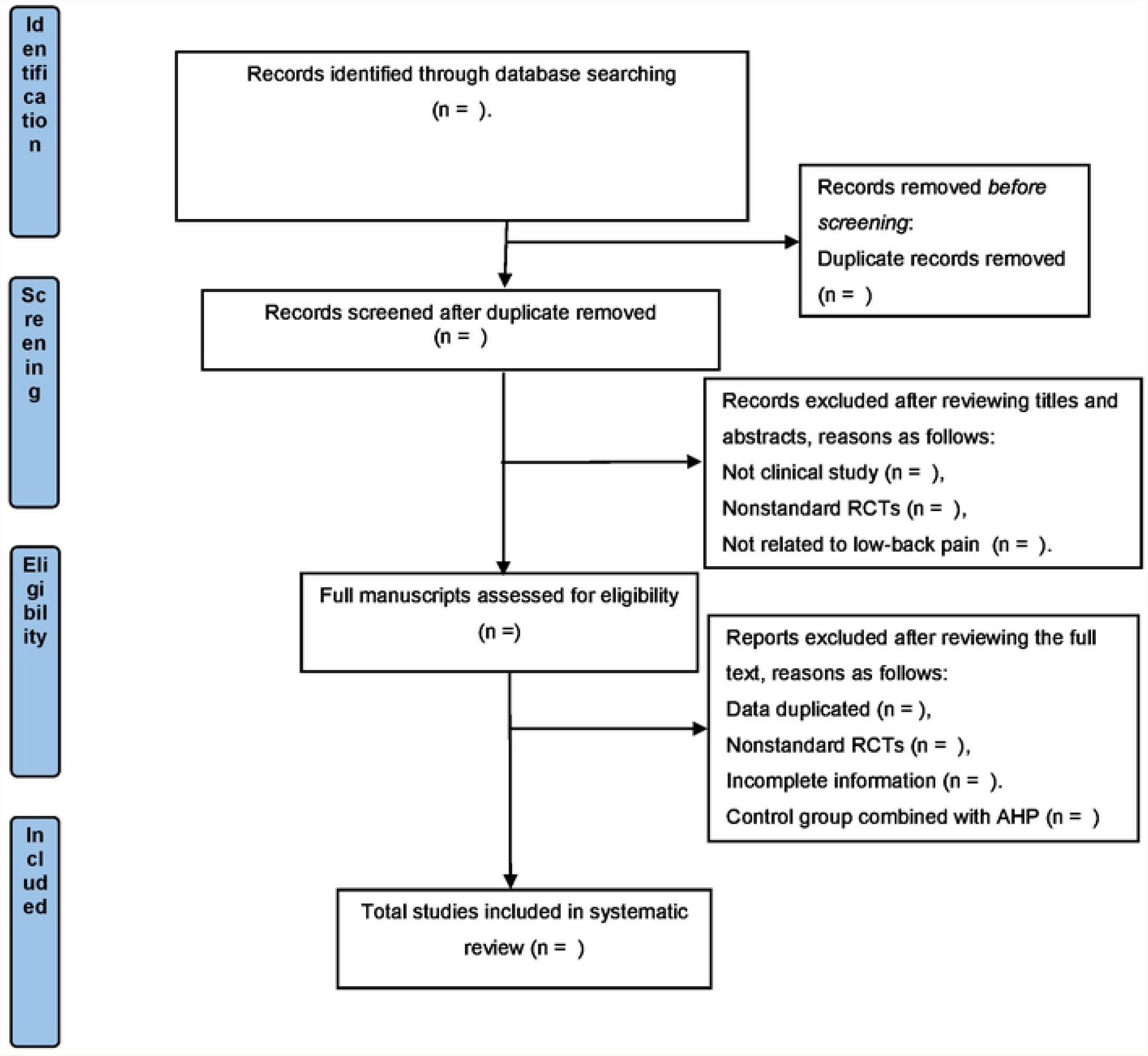
Flow chart of the search process.

## 6. Addressing missing data or unclear measurement scales

We will obtain the missing data or additional information by contacting the study authors via email or telephone if possible.Otherwise, we will analyze the available information and conduct sensitivity analysis to explore the potential impact of insufficient information on the results of the meta-analysis.

## 7. Risk of bias in included studies

Review authors (Ma TS) will independently evaluate each included study and will follow the domain-based evaluation as developed by the Cochrane Handbook for Systematic Reviews of Interventions. They will assess the following domains:

1. selection bias (random sequence generation and allocation concealment),
2. performance bias (blinding of participants and personnel),
3. detection bias (blinding of outcome assessment),
4. attrition bias (incomplete outcome data),
5. reporting bias (selective reporting)
6. other bias (such as pre-sample size estimation, early stop of trial).

Each domain will be divided into 3 categories: “low risk”, “high risk”, or “unclear risk”.

## 8. Data synthesis and analysis

We will analyze the data with RevMan software (Version 5.3)(Available at: https://community.cochrane.org/help/tools-and-software/revman-5) provided by The Cochrane Collabora-tion[27].A meta-analysis using random or fixed effects models will be conducted to summarize the data. Continuous data will be pooled and presented as mean differences or standardized mean difference with their 95% CI. Dichotomous data will be pooled and expressed as risk ratio with their 95% CI. We will interpret it using the following criteria: *I* ^2^ values of 25% is considered low levels of heterogeneity, 50% indicated moderate levels, and 75% indicated high levels[28].Since low or moderate heterogeneity suggests little variability among these studies, the data will be analyzed in a fifixed-effects model[29].When signifificant heterogeneity occurs among the studies (P<.05, I2>50%), a random effect model will be performed to analyze the data.

## 9. Additional analyses

Subgroup analysis will be conducted to evaluate the specific influence of intervention type, age, course of disease, treatment duration on pooled results. If the data is insufficient, qualitative synthesis will be conducted instead of quantitative synthesis. In addition, sensitivity analysis will be performed to examine the robustness of the results by eliminating low quality trials. We willalso use Spass software (Version19.0)

(Available at:https://www.ibm.com/analytics/spss-statistics-software) for complex network analysis to explore the potential core prescription of acupoint herbal patching for NS-LBP.

## 10. Assessment of reporting biases

Reporting bias will be evaluated by visual inspection of Funnel plots. At the same time, Begg test and Egger test will be used to test whether the funnel plot is symmetrical. A P value < 0.05 in Egger test or Begg test is considered statistically significant.

## 11. Confifidence in cumulative evidence

In order to better prepare results for usage in guideline development, We will use the GRADE approach to assess the overall quality of evidence supporting the primary outcomes[30] .GRADE will be used to summarize the limitations in design, consistency, directness, precision, publication bias. The quality of each evidence will be divided into 4 levels: high, medium, low, and very low. Disagreements will be resolved by consensus.

## 12. Discussion

Low back pain (LBP) ranks as the number one disorder in terms of years lived with disability;estimated global one year incidences range from 22% to 65%[31]. Back pain as a symptom, not attributable to spinal instability caused by trauma, infection, progressive deformity, or tumour, and not associated with radicular symptoms, is labelled NS-LBP. LBP is related to disability and work absence and accounts for high economical costs in Western societies [32].Such problems place a great economic burden on society.People with LBP require long-term treatment.The nature of low back pain could also result in less quantifiable costs such as difficulties doing domestic chores, caregiving, engaging in recreational activities, struggles with relationships, depression, and anxiety[33].

At present, the treatment of LBP varies from medical methods. For refractory low back pain, a wide range of non-surgical (eg, epidural steroid injections and spinal cord stimulation for neuropathic pain, and radiofrequency ablation and intra-articular steroid injections for mechanical pain) and surgical (eg, decompression for neuropathic pain, disc replacement, and fusion for mechanical causes) treatment options are available in carefully selected patients. Most treatment options address only single, solitary causes and given the complex nature of low back pain, a multimodal interdisciplinary approach is necessary.AHP as an alternative replacement therapy, more and more accepted by the public and recognition.AHP is a herbal patch that is applied to specific acupoints to stimulate the skin, meridians, and collaterals to produce preventive and therapeutic effects. A study has proved that traditional Chinese medicine can be absorbed from the skin.Studies have also shown that after 4 to 6 hours, the transdermal absorption rate of herbs applied through herbal patches can reach 18.02% to 19.97%[34].The herbal component of the patch contains Analgesic, counter substance(quercetin and Kaempferol).Quercetin can control inflammation factors and inflammatory response, protect oppressed nerve, improvement of morning stiffness and pain[35].Resurrectionlily phenol by blocking MAPK activation pathways, inhibiting inflammatory fiber cell migration, invasion, which block the happening of the pain[36].AHP in medicine include: Chinese ephedra and radix angelicae tuhuo.These two kinds of traditional Chinese medicine (TCM) contains:quercetin and resurrectionlily phenol.Therefore, AHP is gradually applied to the treatment of LBP.There is no planned or published systematic review of the effectiveness and safety of AHP for LBP.The purpose of this study is to evaluate the effect of AHP on clinical effectiveness, functional outcomes, quality of life, improvement of clinical symptoms of LBP, adverse events, and drug withdrawal events in LBP patients. In particular, we will analyze specific acupuncture points and herbal prescriptions used in LBP with Spass software (Version19.0).Herein, this study will be the first to evaluate the clinical efficacy and effective prescription of AHP for LBP patients, and may benefit practitioners in the field of complementary and alternative therapies.

## Data Availability

All relevant data from this study will be made available upon study completion.

## 13. Ethics and dissemination

Ethics approval is not required due to this work is carried out on published data. We aimed to explore the clinical effective rate,functional outcomes, quality of life, improvement of clinical symptoms of LBP, as well as effective prescriptions of AHP for patients with functional constipation.In the end, the results will be submitted to a peer-reviewed journal.

INPLASY registration number: INPLASY202210040

## Author contributions

Conceptualization: Tianshu Ma, Wu Liu.

Data curation: Yan Dong, Xiaona Liu.

Formal analysis: Tianshu Ma,Lei Cheng.

Funding acquisition: Chengyu Liu.

Investigation: Ziyuan Wang.

Methodology:Tianshu Ma, Chengyu Liu.

Project administration: Yan Dong, Li Tie.

Resources: Tianshu Ma,Wu Liu, Chengyu Liu.

Software: Tianshu Ma,Wu Liu.

Supervision:Chengyu Liu.

Validation: Lei Cheng, Xiaona Liu.

Visualization: Chengyu Liu.

Writing-original draft:Tianshu Ma, Wu Liu.

Writing-review & editing: Chengyu Liu, Li Tie.

## Footnotes

We appreciate the financial support received from the Jilin province department of project (grant number: 20210402014GH).

The authors report no conflicts of interest.

The datasets generated during and/or analyzed during the current study are not publicly available, but are available from the corresponding author on reasonable request.

## Abbreviations

AHP: acupoint herbal application
CBM: the Chinese Biomedical Literature Database
LBP: low back pain
NS-LBP: non-specific low back pain
VAS: visual analog scale
NRS: numerical rating scale
CBM: Biomedical Literature Database
NSAIDs: non-steroid anti-inflammatory drugs
RCTs: randomized clinical trials
CI: confidence interval
GRADE: Grading of Recommendations Assessment, Development, and Evaluation
TCM: traditional Chinese medicine

